# Inferring the differences in incubation-period and generation-interval distributions of the Delta and Omicron variants of SARS-CoV-2

**DOI:** 10.1101/2022.07.02.22277186

**Authors:** Sang Woo Park, Kaiyuan Sun, Sam Abbott, Ron Sender, Yinon Bar-on, Joshua S. Weitz, Sebastian Funk, Bryan T. Grenfell, Jantien A Backer, Jacco Wallinga, Cecile Viboud, Jonathan Dushoff

## Abstract

Estimating the differences in the incubation-period, serial-interval, and generation-interval distributions of SARS-CoV-2 variants is critical to understanding their transmission and control. However, the impact of epidemic dynamics is often neglected in estimating the timing of infection and transmission—for example, when an epidemic is growing exponentially, a cohort of infected individuals who developed symptoms at the same time are more likely to have been infected recently. Here, we re-analyze incubation-period and serial-interval data describing transmissions of the Delta and Omicron variants from the Netherlands at the end of December 2021. Previous analysis of the same data set reported shorter mean observed incubation period (3.2 days vs 4.4 days) and serial interval (3.5 days vs 4.1 days) for the Omicron variant, but the number of infections caused by the Delta variant decreased during this period as the number of Omicron infections increased. When we account for growth-rate differences of two variants during the study period, we estimate similar mean incubation periods (3.8–4.5 days) for both variants but a shorter mean generation interval for the Omicron variant (3.0 days; 95% CI: 2.7–3.2 days) than for the Delta variant (3.8 days; 95% CI: 3.7–4.0 days). We further note that the differences in estimated generation intervals may be driven by the “network effect”—higher effective transmissibility of the Omicron variant can cause faster susceptible depletion among contact networks, which in turn prevents late transmission (therefore shortening realized generation intervals). Using up-to-date generation-interval distributions is critical to accurately estimating the reproduction advantage of the Omicron variant.

**Significance:** Recent studies suggest that individuals infected with the Omicron variant develop symptoms earlier (shorter incubation period) and transmit faster (shorter generation interval) than those infected with the Delta variant. However, these studies typically neglect population-level effects: when an epidemic is growing, a greater proportion of current cases were infected recently, biasing us toward observing faster transmission events. Accounting for this dynamical bias, we find that Omicron infections from the Netherlands at the end of December 2021 had similar incubation periods, but shorter generation intervals, compared to Delta infections from the same period. Shorter generation intervals of the Omicron variant might be due to its higher effective reproduction number, which can cause faster local susceptible depletion around the contact network.

## 1 Introduction

Estimating transmission advantages of new SARS-CoV-2 variants is critical to predicting and controlling the course of the COVID-19 pandemic [1]. Transmission advantages of invading variants are typically characterized by the ratios of reproduction numbers, ℛ_inv_/ℛ_res_, and the differences in growth rates, *r*_inv_ − *r*_res_. These quantities are linked by the generation-interval distributions of the resident and invading variants. For example, an invading variant with shorter generation intervals—defined as the time between infection of the infector and the infectee—will exhibit faster epidemic growth (*r*_inv_ *> r*_res_ > 0) even if their reproduction numbers are identical (ℛ_inv_ = ℛ_res_ > 1).

Estimating the generation-interval distribution is challenging, in part due to difficulties in observing actual infection events. Many researchers primarily focus on comparisons of other transmission intervals, such as the time between symptom onsets (also referred to as serial intervals) or between testing events [2] of the infector and the infectee. Each of these transmission-interval distributions can be subject to dynamical effects, which can cause transmission-interval distributions to systematically differ from the corresponding generation-interval distribution.

For example, when the epidemic is growing, there will be more recent infections, and we are therefore more likely to observe recently infected individuals among a cohort of infectors who developed symptoms at the same time. In this case, their incubation periods will be shorter, on average, than those of their infectees, causing the mean serial interval to be longer than the mean generation interval [3]. We refer to such effects of growth rate on expected intervals as “dynamical bias” Because of dynamical bias, observed differences in transmission-interval distributions between variants are not necessarily equivalent to differences in the underlying generation-interval distributions when their growth rates differ.

Here, we re-analyze serial-interval data collected by [4], representing within- and between-household transmissions of the B.1.617.2 (Delta) and B.1.1.529 (Omicron) variants from the Netherlands between 13 and 26 December 2021. The study found shorter mean within-household serial intervals (3.5 vs 4.1 days) and mean incubation periods (3.2 vs 4.4 days) for transmission pairs with S-gene target failure (mostly Omicron during the study period) than without (mostly Delta), but did not consider dynamical biases caused by growth-rate differences in their inference: during this period, the incidence of Omicron cases were increasing, whereas the incidence of Delta cases were decreasing. We take the epidemiological context in the Netherlands during the study period into account to provide corrected estimates for the incubation periods and generation-interval distributions of the Delta and Omicron variants. We show that using up-to-date generation-interval distributions is critical to accurately estimating the reproduction advantage (i.e., the ratio between the reproduction numbers of the invading and resident variants) of emerging SARS-CoV-2 variants.

## 2 Methods

### 2.1 Data

We analyze time series of reported COVID-19 cases (https://data.rivm.nl/covid-19/) and proportions of SARS-CoV-2 variants detected (https://www.rivm.nl/coronavirus-covid-19/virus/varianten) from the Netherlands between 29 November 2021 and 30 January 2022. Data sets are publicly available on the National Institute for Public Health and the Environment (RIVM) website.

Serial-interval data are taken from [4]. Infector-infectee pairs were identified through contact tracing, and their symptom onset dates were reported through a national surveillance database. Serial intervals were then calculated by taking the difference between symptom onset dates of the infector and the infectee. In order to ensure independence between serial intervals, one infectee was chosen at random for each infector in the original analysis. See original article for additional details of data collection.

Publicly available data are aggregated by the length of the serial interval in days and do not include additional individual-level information, such as exposure dates, symptom onset dates, or age. The original article presented serial-interval estimates stratified by the vaccination status in supplementary materials, but stratified data are not publicly available; we rely on publicly available data to keep the analysis simple and to focus on the qualitative effects of dynamical biases. The aggregated data consists of 2529 transmission pairs and are further stratified by the presence of S gene target failure (SGTF), week of infectors’ symptom onset date (week 50, 13–19 December 2021, and week 51, 20–26 December 2021), and the type of transmission (within- or between-household transmission). In the main text, we combine data from weeks 50 and 51 of 2021 (13–26 December) and present a stratified analysis in Supplementary Material. For simplicity, we refer to transmission pairs with and without SGTFs as Omicron and Delta transmission pairs, respectively. Incubation period data were originally collected from 513 individuals (consisting of 258 Omicron and 255 Delta cases), with symptom onsets between 1 December 2021 and 2 January 2022; however, the data are not publicly available with the original article. Instead, we rely on previous estimates [4] to derive growth-rate-adjusted incubation-period distributions.

### 2.2 Estimating epidemic growth rates

In order to accurately estimate incubation-period and generation-interval distributions of the Delta and Omicron variants, we have to take their epidemiological dynamics—in particular, their growth rates—into account. To estimate the growth rates of the Delta and Omicron variants, we first estimate the number of COVID-19 cases infected with each variant by multiplying reported weekly numbers of cases by the proportion of Delta and Omicron variants detected—we use weekly time series to smooth over patterns of testing and reporting within each week. We note that the proportion of Delta and Omicron variants detected is reported with the date of sampling, whereas the case data are reported with the date of report, meaning that there is some delay between the two data sets (typically around 2 days). For simplicity, we do not account for this delay in our growth-rate estimates; instead, we later perform sensitivity analysis to assess how growth rates affect the inferences of the incubation-period and generation-interval distributions. We also do not account for uncertainties around the estimates of the proportion of each variant—almost 2000 samples were tested on each week between the week of November 28, 2021, and the week of January 23, 2022, making uncertainty due to sample size small; we note however that this estimate is also sensitive to the assumption that sampling is random.

We then fit a generalized additive model [5] to the logged weekly case estimates to obtain smooth trajectories for case time series. More specifically, we model the logged weekly numbers of cases infected with each variant as a function of time using a penalized cubic spline fitted with restricted maximum likelihood (specified as `gam(log(cases) ∼s(time, bs=“cs”), method=“REML”)` using the `MGCV R` package): We use Gaussian likelihoods to fit to logged cases in part for convenience, and in part because the inferred numbers of cases infected with each variant are not whole numbers. In principle, it might be preferable to explicitly model the process of sampling for strain testing, and then using negative-binomial likelihoods for case numbers [6], but our main purpose here is simply to roughly estimate growth rates with reasonable uncertainties. Finally, we take the derivative of the predicted logged numbers of cases infected with each variant to obtain time-varying growth rate estimates.

To obtain confidence intervals on the estimated time-varying growth rates, we generate 1000 parameter sets by resampling spline coefficients from a multivariate normal distribution using the estimated variance-covariance matrices. We calculate time-varying growth rates from each parameter set and use equi-tailed quantiles to generate 95% confidence limits. We note that this method of calculating confidence intervals gives point-wise confidence intervals, meaning that the confidence intervals give 95% coverage for the set of estimates at each time point; these intervals are narrower than simultaneous confidence intervals, which give 95% coverage for the set of estimated time series across the whole time period [7].

### 2.3 Estimating forward incubation-period distributions from backward incubation-period distributions

The incubation-period distributions from 513 individuals (consisting of 258 Omicron and 255 Delta cases), with symptom onsets between 1 December 2021 and 2 January 2022, were previously reported in [4]. These data cover a wider time period than the serial-interval data. [4] used the methods of [8], which estimates incubation period by inferring distributions of time of infection for each individual from their known exposure dates. In particular, the methods of [8] assume that the infection time is uniformly distributed across exposure dates and compares the inferred infection time to a known symptom-onset time to calculate the incubation period for each individual. Even if this method can accurately estimate the infection time, and therefore the incubation period, of each individual, dynamical biases can still affect this sort of cohort-based estimation of incubation period.

More specifically, incubation periods (and other epidemiological delays) can be measured in two ways: forward and backward [3]. Forward incubation periods are measured from a cohort of individuals who were infected at the same time. We expect the forward incubation-period distribution *f*_*I*_(*τ*) to remain relatively constant over the course of an epidemic of one given variant, although biases can arise in *observing* incubation periods, based on public or medical awareness of the disease. Backward incubation periods are measured from a cohort of individuals who developed symptoms at the same time. The backward incubation-period distribution is sensitive to epidemic dynamics: the difference between the forward and backward distribution arises because forward incubation periods look forward from the reference point towards symptom development, which is an individual-level process, while backward incubation periods look backwards towards an infection event, which requires an interaction with an infectious individual.

In particular, when incidence of infection is growing exponentially, we are more likely to observe backward incubation periods that are shorter than the corresponding forward incubation periods because there will be relatively more individuals who were infected recently. Assuming that incidence of infection is changing exponentially at a constant rate *r* across the study period, the backward incubation-period distribution *b*_*I*_(*τ*) corresponds to:

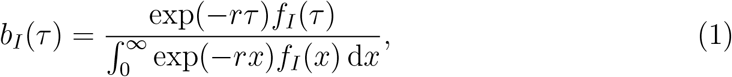

where the denominator is a normalization constant so that *b*_*I*_(*τ*) integrates to 1. Therefore, the backward incubation-period distribution *b*_*I*_(*τ*) gives a biased estimate of the corresponding forward distribution *f*_*I*_(*τ*). The method of [8] starts from observed symptom onsets, and estimates the backward incubation-period distribution.

Assuming a constant growth rate *r*, the corresponding forward incubation-period distributions can be calculated by inverting Eq. (1), taking into account that *f*_*I*_ is a probability distribution and therefore needs to be normalised to integrate to 1:

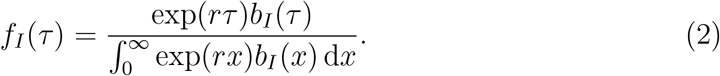

Since incubation-period data are not provided, we are not able to fit Eq. (2) directly; instead we take the backward incubation-period distributions *b*_*I*_(*x*) estimated by [4], which was originally assumed to follow a Weibull distribution, and apply Eq. (2). In particular, [4] estimated the scale and shape parameters of the Weibull distribution to be 4.93 (95% CI: 4.51–5.37) and 1.83 (95% CI: 1.59–2.08), respectively, for the Delta cases, and 3.60 (95% CI: 3.23–3.98) and 1.50 ((95% CI: 1.32–1.70), respectively, for Omicron cases.

We also model the backward incubation-period distribution *b*_*I*_(*τ*) using a Weibull distribution based on the assumptions of [4]. To account for uncertainties in the original parameter estimates, we rely on a sampling scheme, similar to the one we used for the growth rate analysis (in Section 2.2). First, we approximate the previously inferred posterior distributions of the shape and scale parameters of the Weibull distribution using a lognormal distribution—we parameterize the lognormal distribution such that (i) its median matches the median of the posterior distributions and (ii) the probability that a random variable following the specified lognormal distribution falls between the lower and upper credible limits is 95% [9]. We draw 1000 samples of the shape and scale parameters (for the backward distribution *b*_*I*_(*τ*)) from the specified lognormal distributions and estimate the corresponding forward distribution using Eq. (2). We take 95% equi-tailed quantiles to obtain 95% confidence intervals. We repeat the analysis across plausible ranges of *r* for the Delta and Omicron variants separately (discussed later).

### 2.4 Estimating forward generation-interval distributions from forward serial-interval distributions

Dynamical biases in the serial-interval distributions are more complex because the serial interval depends on the incubation periods of the infector and the infectee as well as the generation interval between them (Fig. 1). For example, [4] measured the forward serial-interval distributions from cohorts of infectors who developed symptoms during the same week. In this case, the forward serial interval *τ*_*s*_ can be expressed in the form [3]:

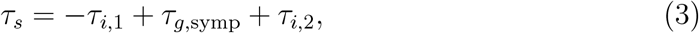

where *τ*_*i*,1_ represents the backward incubation period of the infector (because all infectors developed symptoms at the same time), and *τ*_*i*,2_, represents the forward incubation period of the infectee. Here, *τ*_*g*,symp_ represents the generation interval between the infector and the infectee; we use the subscript symp to indicate that these generation intervals are measured from infectors who developed symptoms at the same time.

**Figure 1:**
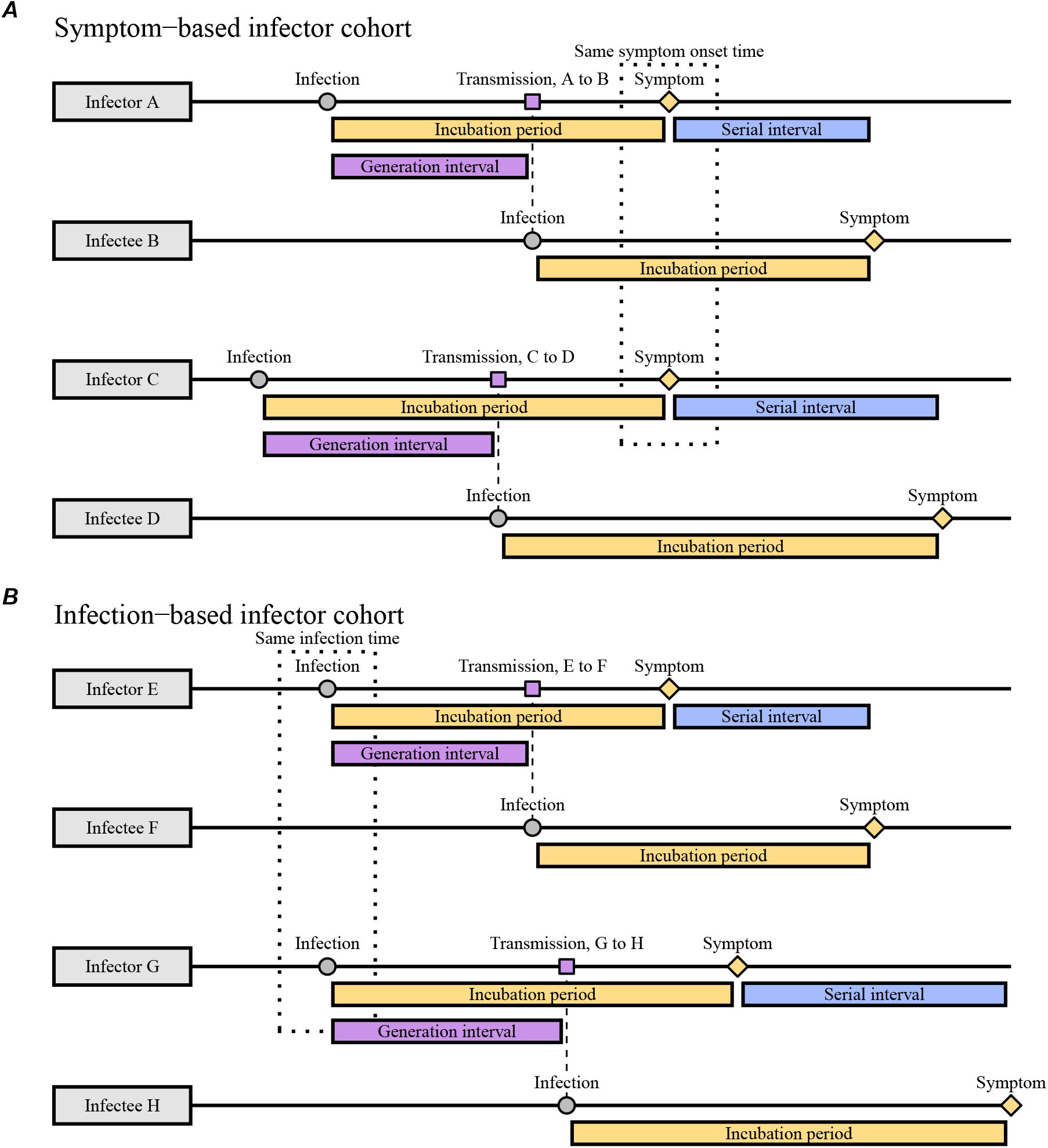
Schematic diagrams of serial and generation intervals from symptom- and infection-based infector cohorts. (A) Forward serial intervals are measured from the cohort of infectors who develop symptoms at the same time. In this case, infectors will have shorter incubation periods than their infectees on average; the corresponding generation intervals will be also short because infectors with short incubation periods will transmit earlier. (B) Generation intervals for the cohort of infectors who are infected at the same time are not biased by dynamical effects on incubation periods.

The generation-interval distribution for a symptom-based cohort (*τ*_*g*,symp_ in Eq. (3)) is biased (compared to the generation-interval distribution for an infection-based cohort) because infectors who developed symptoms at the same time will have shorter incubation periods (when the epidemic is growing) and are therefore likely to transmit earlier (Fig. 1A). This generation-interval distribution for a symptom-based cohort depends on the backward incubation-period distribution:

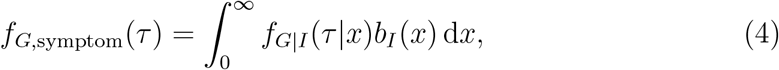

where *f*_*G*|*I*_ (*τ*|*x*) represents the forward generation-interval distribution conditional on a known value of the incubation period, *x*, and *b*_*I*_(*x*) represents the backward incubation-period distribution. Instead, the forward generation-interval distribution measured from a cohort of individuals who were infected at the time is expected to provide reliable estimates of the distribution across individuals (because their incubation-period distribution is expected to remain constant over time, Fig. 1B):

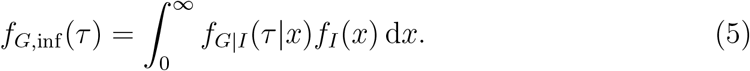

Previous analyses of serial-interval distributions typically assumed that the incubation periods and generation intervals are independent [10]; in this case, the generation-interval distribution for the symptom-based and infection-based cohorts are identical. In summary, when an epidemic is growing exponentially, there are two opposing effects affecting the relationship between the mean forward serial and generation interval. First, infectors who developed symptoms at the same time are more likely to have shorter (backward) incubation periods than the corresponding forward incubation periods of their infectees on average, 𝔼[*τ*_*i*,1_] < 𝔼[*τ*_*i*,2_], causing the mean forward serial interval to be longer than the mean symptom-based generation interval (𝔼[*τ*_*s*_] > 𝔼[*τ*_*g*,symp_]). Second, the mean symptom-based generation interval will be shorter than the mean infection-based generation interval: *E*[*τ*_*g*,inf_] > 𝔼[*τ*_*g*,symp_] due to correlations between incubation periods and generation intervals. Therefore, the difference between the mean forward serial interval and the mean infection-based generation interval is difficult to predict in general; in most cases, however, we expect the former effect to dominate, causing the mean forward serial interval to be longer than the mean infection-based generation interval: 𝔼[*τ*_*s*_] > *E*[*τ*_*g*,inf_] [3]. Earlier work on serial-interval distributions neglected dynamical biases in the incubation periods of the infectors [11, 12], which allowed the authors to conclude that the mean generation and serial intervals are identical. For simplicity, we will use the term “forward generation-interval” to refer to the infection-based generation-interval distribution (measured from a cohort of infectors who were infected at the same infection time, Fig. 1B), and drop the subscript inf.

Assuming that the incidence of infection will continue to change exponentially at a constant rate *r*, the forward serial-interval distribution for a cohort of infectors who developed symptoms at the same time *t* is expected to remain unchanged through time [3]. Then, we can focus on the forward serial-interval distribution at *t* = 0, which in turn allows us to reparameterize the incubation-period and generation-interval distributions in terms of the infection time of the infector *α*_1_ *<* 0 and of the infectee *α*_2_ *> α*_1_. Under this parameterization, for a given length of a serial interval *τ*, we can rewrite the incubation period of the infector as −*α*_1_; the generation interval as *α*_2_ − *α*_1_; and the incubation period of the infectee as *τ* − *α*_2_. Then, the forward serial-interval distribution *f*_*S*_(*τ*) for a cohort of infectors who developed symptoms at time *t* = 0 can be expressed in terms of three distributions (Eq. (3)): the backward incubation-period distribution of the infector *b*_*I*_(−*α*_1_) (taken from Eq. (1)), the forward generation-interval distribution conditional on a known value of the incubation period *x, f*_*G*|*I*_ (*α*_2_ − *α*_1_| − *α*_1_), and the forward incubation-period distribution of the infectee *f*_*I*_(*τ* − *α*_2_). Integrating across infection time of the infector *α*_1_ *<* 0 and of the infectee *α*_2_ *> α*_1_ and rewriting the backward incubation-period distribution *b*_*I*_(−*α*_1_) in terms of the forward distribution, we obtain [3]:

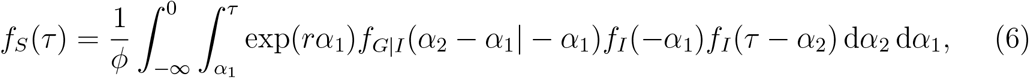

where *ϕ* is a normalization constant chosen so that *∫ f*_*S*_(*x*) d*x* = 1. As discussed earlier, this method assumes that the incidence is changing exponentially at a constant rate *r* across the study period. As we show in Results, the exponential growth rate changes over the study period, including weeks 50 and 51 (13–26 December 2021); for illustrative purposes, we choose representative values of *r* that for Delta and Omicron during this period and also explore across plausible ranges of *r* (see below).

While the derivation of the forward serial-interval distribution Eq. (6) may be complex, its implementation is simple. The main difference between our model and previous models that neglect dynamical effects [10, 13, 14, 15] is the exponential growth term exp(*rα*_1_) and the normalization term *ϕ*—it is relatively straightforward to include these terms in existing models of serial intervals. [16, 17] also included this term in their analyses of serial-interval data, but only accounted for the epidemic growth effect (and not the decay effect).

We model the forward incubation-period *f*_*I*_(*τ*) and generation-interval *f*_*G*_(*τ*) distributions using a bivariate lognormal distribution. The joint distribution is parameterized by log-scale means, *μ*_*I*_ and *μ*_*G*_, log-scale variances, 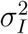 and 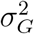, and the log-scale correlation coefficient *ρ*. Thus, the forward generation-interval distribution conditional on the incubation period *f*_*G*|*I*_ (*τ*|*τ*_*i*,1_) has a log-scale mean of *μ*_*G*_ + *σ*_*G*_*ρ*(log(*τ*_*i*,1_) − *μ*_*I*_)*/σ*_*I*_ and a log-scale variance of 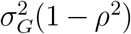. For a given value of *r*, we first estimate the forward incubation-period distribution from the backward distribution, previously estimated by [4], using Eq. (2). We then approximate the forward incubation-period distribution with a lognormal distribution by matching the mean and standard deviation (also known as the method of moments); we note that we are unable to directly fit a lognormal distribution to the forward incubation-period distribution because we are relying on existing estimates rather than raw data. Using this incubation-period distribution, we fit Eq. (6) to the observed serial-interval data by minimizing the negative log-likelihood. We then calculate the mean forward generation interval using Eq. (5). The 95% confidence intervals are calculated by taking the estimated variance-covariance matrix for our mean and standard deviation parameters and calculating the corresponding variance-covariance for the overall mean using Taylor expansion—this method is also known as the Delta method [18]. We assume *ρ* = 0.75 throughout based on [19]—since we do not have individual-level data on infection and symptom onset times, we expect this parameter to be unidentifiable in practice. In Supplementary Material, we explore how assumptions about *ρ* affect inferences of the generation-interval distribution.

### 2.5 Estimating instantaneous reproduction numbers

We use our estimates of the generation-interval distributions to infer instantaneous reproduction numbers ℛ(*t*) of the Delta and Omicron variant, as well as the ratio between the two reproduction numbers. Estimating the instantaneous reproduction number—defined as the average number of secondary infections that a primary case will generate if epidemiological conditions remain constant [20]—requires the intrinsic generation-interval distribution *g*(*τ*):

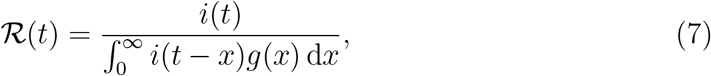

where *i*(*t*) represents incidence of infection. Here, we approximate the intrinsic generation-interval distribution with the forward generation-interval that we estimate for weeks 50 and 51 of 2021 (13–26 December)—when the epidemic is growing or decaying exponentially, we expect the forward generation-interval to be a good proxy for the intrinsic generation-interval distribution [21, 22]. Incidence of infection is approximated by shifting the smoothed case trajectories by one week to account for reporting delays. This method of approximating incidence of infection assumes a fixed delay between infection and case reporting; in practice, deconvolution is required to accurately estimate the incidence of infection [23]. Case reports are also sensitive to changes in testing behavior, and therefore our estimates of ℛ(*t*) must be interpreted with care. Confidence intervals are calculated by sampling parameters of the smoothed case trajectories as well as the generation-interval distributions from multivariate normal distributions and repeating the analysis 1000 times.

## 3 Results

Fig. 2 summarizes the epidemiological context in the Netherlands during the study period. The first known Omicron case in the Netherlands was sampled on 19 November 2021 [4], during a period when COVID-19 incidence was decreasing (Fig. 2A). As the Omicron variant continued to spread and increase in proportion (Fig. 2B), the number of COVID-19 cases started to increase (Fig. 2A). Multiplying the proportion of each variant with the number of reported COVID-19 cases further allows us to estimate the epidemiological dynamics of each (Fig. 2C). The number of COVID-19 cases infected with the Delta variant continued to decrease throughout the study period with time-varying growth rates decreasing from *r* ≈ −0.01/day to *r* ≈ −0.09/day by the week of January 16, 2022, and increasing back up to *r* ≈ −0.04/day by the end of January, 2022 (Fig. 2D). The number of COVID-19 cases infected with the Omicron variant increased rapidly but decelerated over time with time-varying growth rates decreasing from *r* = 0.18/day on the week of December 19, 2021, to *r* = 0.04/day by the end of January, 2022. These changes in growth rates coincide with the introduction of lockdown on 19 December 2021 [24] and its relaxation beginning 15 January 2022 [25, 26]. We note that the growth-rate difference between the Delta and Omicron variants decreased over time. Hereafter, we use *r* = −0.05/day for the Delta variant and *r* = 0.15/day for the Omicron variant as representative growth rates—these growth rates correspond to the mean growth rates between 1 December 2021 and 2 January 2022, during which the incubation-period data were collected. We then evaluate the growth-rate effects across *r* = −0.1/day–0.0/day for the Delta variant and *r* = 0.1/day–0.2/day for the Omicron variant as a sensitivity analysis.

**Figure 2:**
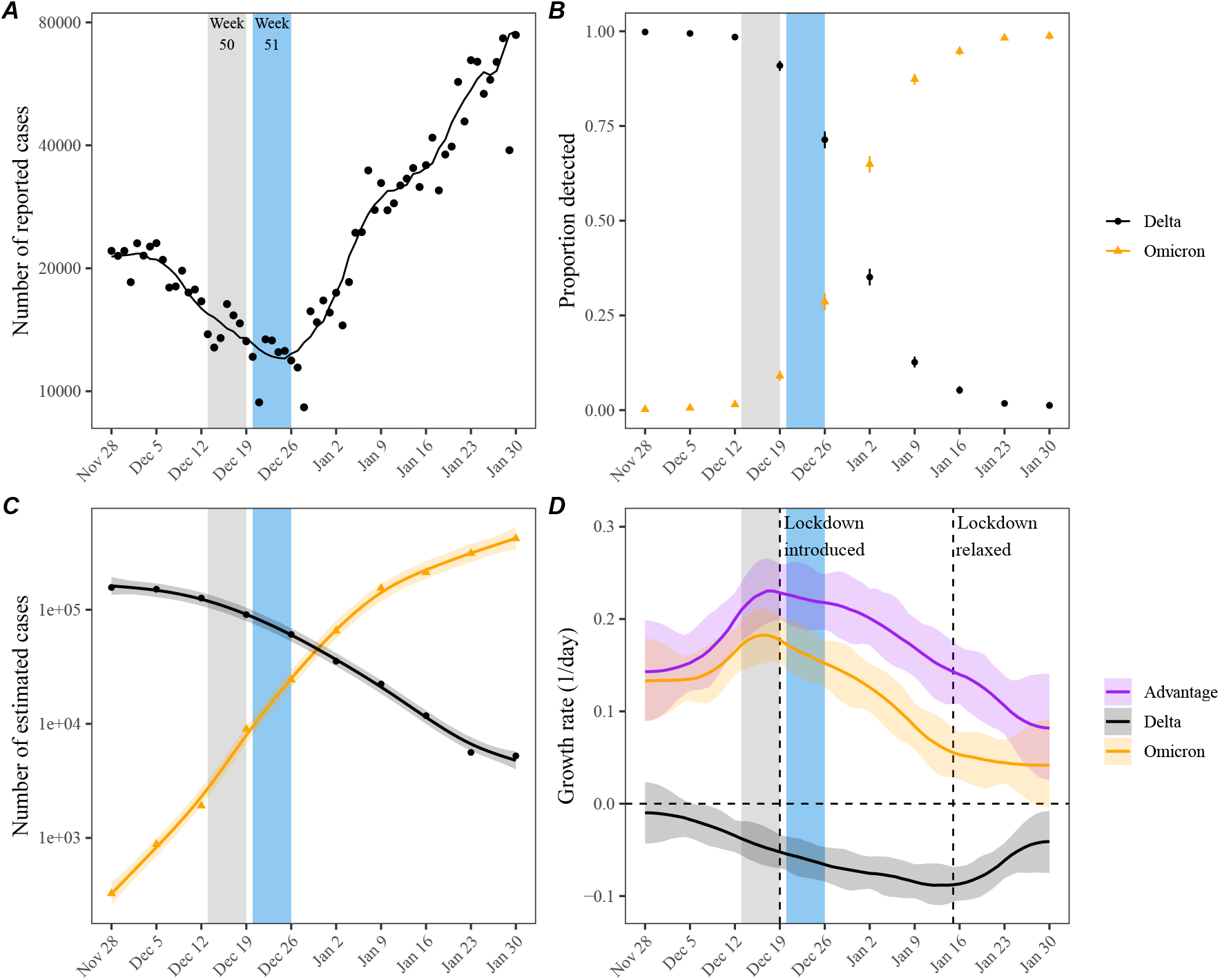
Epidemic dynamics of the Delta and Omicron variants in the Netherlands between November 2021 and January 2022. (A) Daily numbers of reported COVID-19 cases in the Netherlands (points). The solid line represents the 7-day moving average. Data are publicly available at https://data.rivm.nl/covid-19/. (B) Proportion of SARS-CoV-2 variants detected from the Netherlands. Data are publicly available at https://www.rivm.nl/coronavirus-covid-19/virus/varianten. (C) Weekly numbers of COVID-19 cases infected with the Delta (black points) and Omicron (orange triangles) variants are estimated by multiplying the weekly numbers of cases (A) with the proportion of each variant (B). Solid lines and shaded areas represent fitted lines and corresponding 95% confidence intervals using generalized additive model. (D) Estimated growth rates of the Delta (black) and Omicron variants (orange) and their growth-rate differences (purple). Lines and shaded areas represent medians and corresponding 95% confidence intervals. Growth rates are estimated by taking the derivative of the generalized additive model estimates of logged number of cases.

Previous analysis of a cohort of individuals who developed symptoms between 1 December 2021 and 2 January 2022 found longer mean (backward) incubation period for the Delta variant than for the Omicron variant [4] (Fig. 3A). However, when we account for growth-rate differences and re-estimate the forward incubation periods, we find that both variants have similar incubation-period distributions with a mean of 4.1 days (95% CI: 3.8–4.4 days) for the Delta variant and 4.2 days (95% CI: 3.6– 4.9 days) for the Omicron variant Fig. 3B). In this case, the difference between the mean backward and forward incubation periods correspond to −22% and 7% bias for the Omicron and Delta variants, respectively. Although the exact estimate of the mean forward incubation periods of both variants are sensitive to the assumed growth/decay rates, we find similar means across a plausible ranges of growth rates (Fig. 3C–D). For example, the mean forward incubation period of the Delta variant changes from 3.8 days (95% CI: 3.5–4.1 days) to 4.4 days (95% CI: 4.0–4.8 days) as we change the assumed values of *r* from −0.1/days to 0.0/days (Fig. 3C), while the mean forward incubation period of the Omicron variant changes from 3.8 days (95% CI: 3.4–4.4 days) to 4.5 days (95% CI: 3.9–5.5 days) as we change the assumed values of *r* from 0.1/days to 0.2/days (Fig. 3D). Wider confidence intervals for the Omicron variant are driven by greater uncertainties from the dynamical correction, which is larger for Omicron because of higher absolute growth rates.

**Figure 3:**
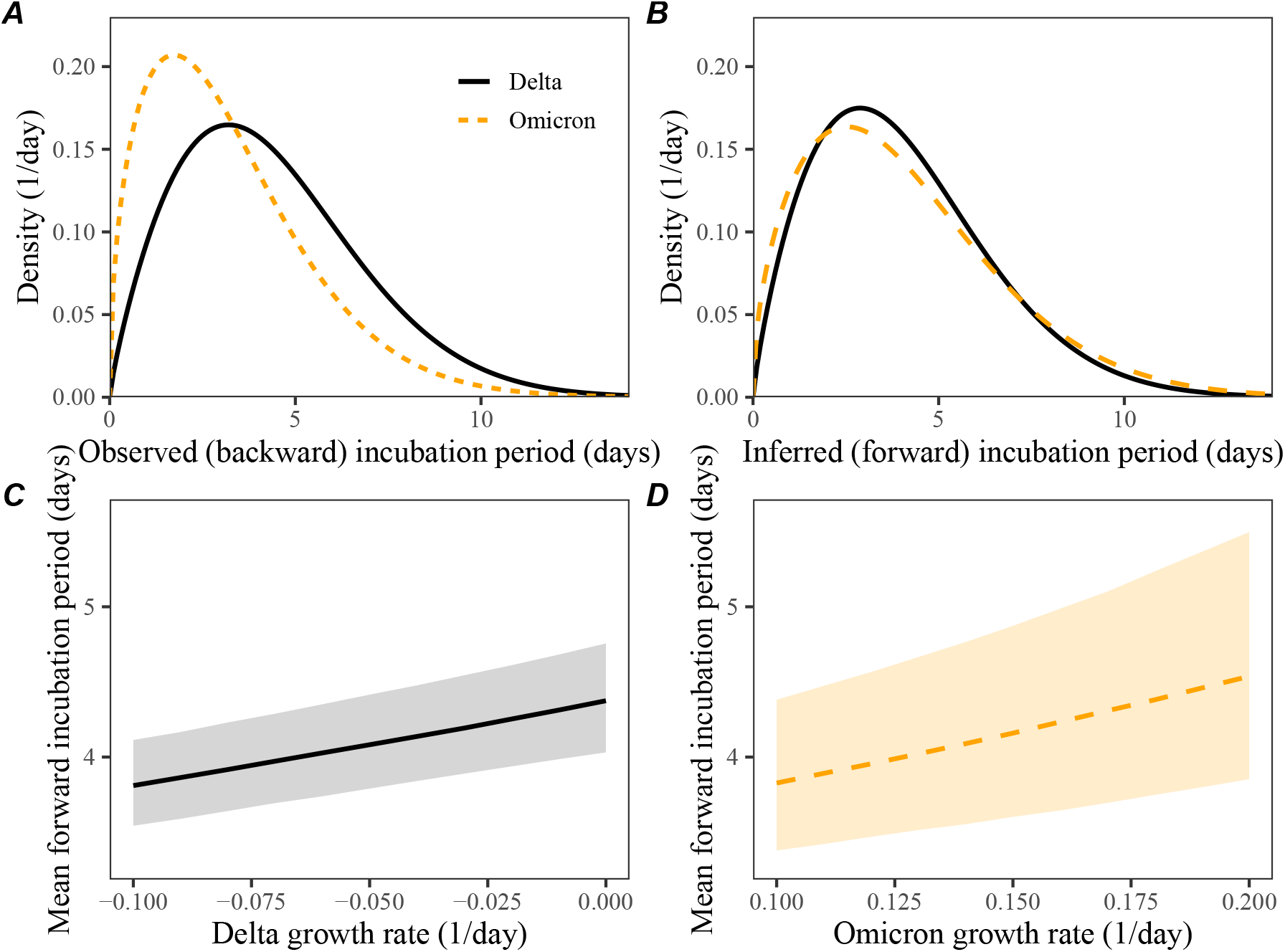
Observed and corrected differences in incubation-period distributions of Delta and Omicron variants. (A) Posterior median estimates of the observed (backward) incubation periods of the Delta (black) and Omicron (orange) variants by [4]. (B) Forward incubation-period distributions assuming *r* = 0.05/day and *r* = −0.15/day for the Delta (black) and Omicron (orange) variants, respectively. (C–D) Corrected estimates of the mean forward incubation-period for different assumptions about the growth rates of the Delta (C) and Omicron variants (D). Lines represent median estimates. Shaded regions represent the corresponding 95% confidence intervals.

We can use these estimates of the forward incubation-period distributions to estimate the forward generation-interval distributions. For illustrative purposes, we first focus on aggregated serial intervals from infectors who developed symptoms during week 50–51 (13–26 December, 2021). For within-household transmission pairs (Fig. 4A), the Omicron variant has shorter mean serial interval (3.1 days; 95% CI: 2.9–3.3 days) than that of the Delta variant (3.7 days; 95% CI: 3.5–3.8 days). When we account for growth-rate differences (assuming *r* = −0.05/day and *r* = 0.15/day for the Delta and Omicron variants, respectively), the estimated mean forward generation interval exhibits a slightly larger difference (Fig. 4B): 3.0 days (95% CI: 2.7–3.2 days) for the Omicron variant and 3.8 days (95% CI: 3.7–4.0 days) for the Delta variant. Our estimate of this difference in these mean generation intervals is robust across plausible ranges of assumptions about the growth rates of the variants (Fig. 4C–D). Assuming lower values of the correlation *ρ* between the incubation period and generation intervals leads to larger differences in the mean generation intervals of the Delta and Omicron variants (Supplementary Figure S1). In particular, the generation-interval estimates of the Omicron variant are more sensitive to the assumed values of *ρ* due to faster changes in incidence of infection—for example, changing *ρ* from 0.85 to 0.5 changes the mean generation-interval estimates for the Omicron variant from 3.1 days (95% CI: 2.8–3.3 days) to 2.7 days (95% CI: 2.5–2.9 days). We explore a wide range of *ρ* to consider the possibility that our original assumption (*ρ* = 0.75) may under or over-estimate the true *ρ*.

**Figure 4:**
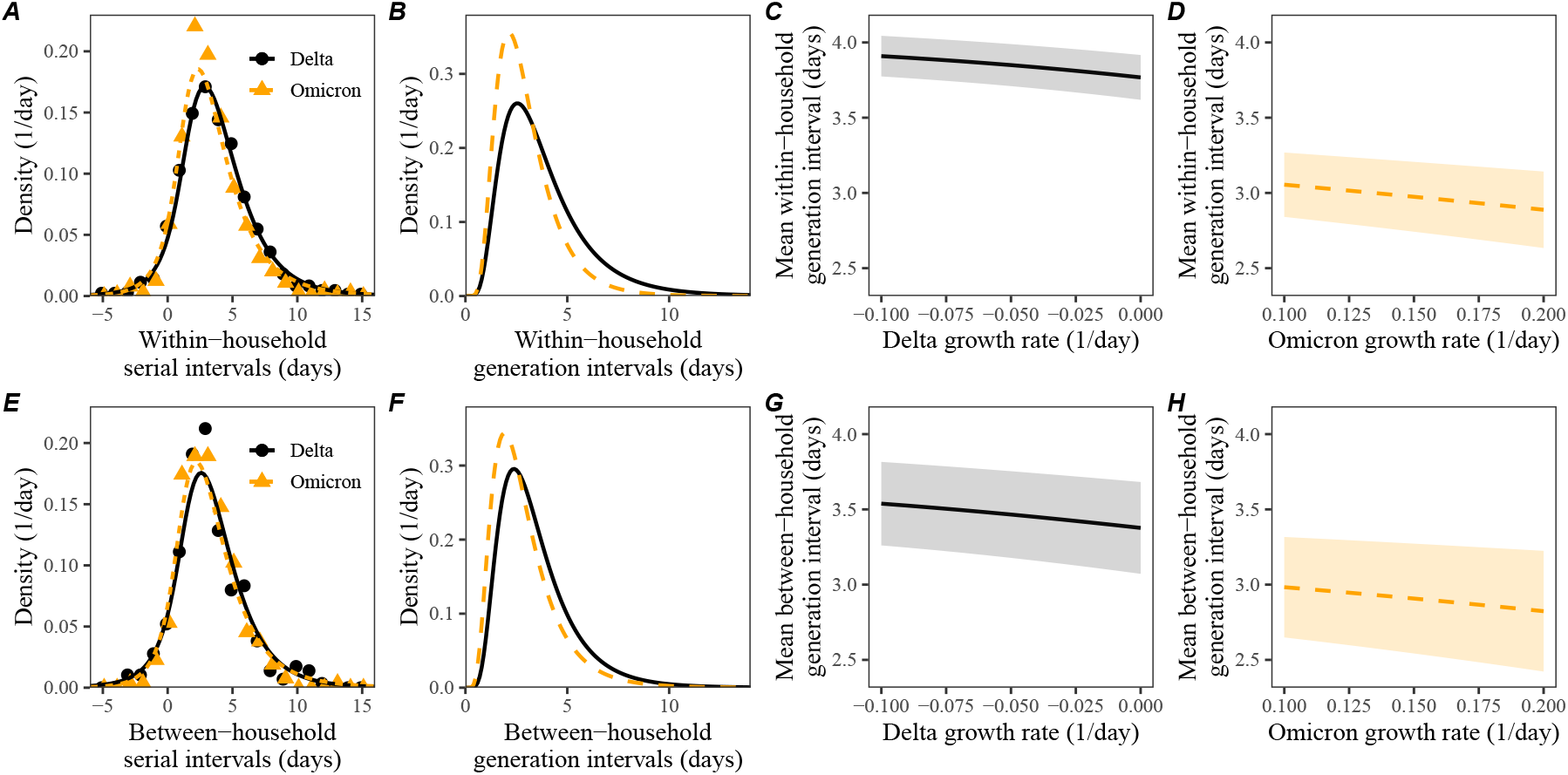
Estimated forward generation-interval distributions of Delta and Omicron variants. (A, E) Observed and fitted forward serial-interval distributions for within-household (A) and between-household (E) transmission pairs in the Netherlands for the Delta (black) and Omicron (orange) variants [4]. Serial intervals are calculated for infectors who developed symptoms on weeks 50 and 51 (13–26 December, 2021). Points represent the observed data. Lines represent the fitted lines assuming *r* = −0.05/day for the Delta variant and *r* = 0.15/day for the Omicron variant. (B, F) Estimated forward generation-interval distributions for within-household (B) and between-household (F) transmission pairs in the Netherlands. (C, D, G, H) Sensitivity of the mean forward generation-interval estimates to assumed growth rates of the Delta (C, G) and Omicron variants (G, H) for within-household (C, D) and between-household (G, H) transmission pairs. Lines represent maximum likelihood estimates. Shaded regions represent the corresponding 95% confidence intervals.

Similar pictures arise for between-household transmission pairs, but the differences in mean serial intervals are unclear (Fig. 4E): 3.0 days (95% CI: 2.7–3.3 days) for the Omicron variant and 3.3 days (95% CI: 3.0 days–3.6 days) for the Delta variant. Consistent with the original study, which also reported shorter mean serial intervals for between-household pairs [4], we estimate shorter mean generation intervals for between-household Delta pairs. While the difference in mean generation intervals is larger, there is greater uncertainty in their mean estimates (Fig. 4F): 2.9 days (95% CI: 2.5–3.3 days) for the Omicron variant and 3.5 days (95% CI: 3.2–3.8 days) for the Delta variant. Once again, these patterns are robust across plausible ranges of assumptions about the growth rates of the Delta and Omicron variants (Fig. 4G–H).

In Supplementary Figure S2, we present generation-interval estimates that are further stratified by the week of infectors’ symptom onset (13–19 December 2021 and 20–26 December 2021). While we generally estimate shorter mean generation intervals for the Omicron variant, but the differences are unclear across all strata, except for within-household transmission pairs during week 50 (13–19 December 2021). We also estimate a reduction in the mean forward generation intervals from week 50 (13–19 December 2021) to week 51 (20–26 December 2021), especially for the Delta variant; this decrease in the mean generation interval is likely associated with the lockdown.

Accounting for differences in the generation-interval distributions, we estimate that the instantaneous reproduction number of the Omicron variant decreased from 1.73 (95% CI: 1.59–1.89) to 1.14 (95% CI: 1.00–1.32) between December 12, 2021, and January 23, 2022(Fig. 5A). On the other hand, the instantaneous reproduction number of the Delta variant decreased from 0.90 (95% CI: 0.83–0.97) to 0.69 (95% CI: 0.65–0.75) between December 5, 2021, and January 9, 2022, and increased back up to 0.83 (95% CI: 0.73–0.94) by January 23, 2022 (Fig. 5A). We estimate the reproduction advantage (i.e., the ratio between the instantaneous reproduction numbers of the Omicron and Delta variants) stayed roughly constant at around 2.10 (95% CI: 1.90–2.33) between December 12–26, 2021, and slowly decreased to 1.38 (95% CI: 1.15–1.65). However, if we neglect differences in the generation-interval distributions and solely rely on the generation-interval-distribution estimate for the Delta variant, we over-estimate the reproduction number of the Omicron variant and therefore the reproduction advantage (Fig. 5B). In this case, the reproduction advantage decreases from 2.38 (95% CI: 2.13–2.67) to 1.43 (95% CI: 1.17–1.75), corresponding to roughly 4–13% bias. Using between-household generation intervals also gives similar conclusions about changes and biases in the reproduction number estimates (Supplementary Figure S3).

**Figure 5:**
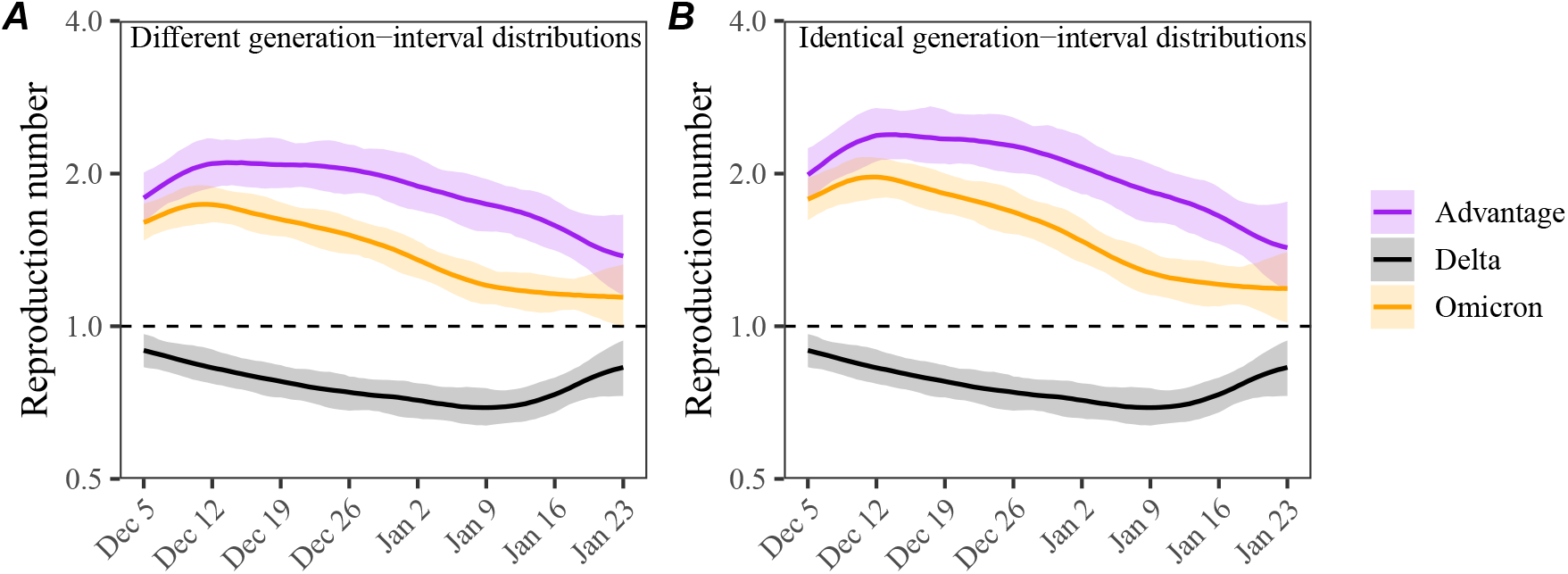
Estimated instantaneous reproduction number advantages of the Omicron variant. (A) Estimated instantaneous reproduction numbers and their ratios over time while accounting for differences in the generation-interval distributions. (B) Estimated instantaneous reproduction numbers and their ratios over time while assuming identical generation-interval distributions. The instantaneous reproduction number of each variant is estimated using the renewal equation by shifting the smoothed case curves by one week (Fig. 2C). The intrinsic generation-interval distribution is approximated by the maximum likelihood estimates of the forward generation-interval distributions for within-household transmission pairs based on *r* = −0.05 for the Delta variant (black) and *r* = 0.15 for the Omicron variant (orange). Purple lines represent the ratio between the effective reproduction numbers of the Delta and Omicron variants. Lines and shaded regions represent medians and corresponding 95% confidence intervals.

In both cases, the decrease in the reproduction advantage coincides with the decrease in the reproduction number of the Omicron variant, implying that epidemiological changes driving the dynamic had larger effects on the transmission of the Omicron variant than on the transmission of Delta variant; a larger reduction in the reproduction number of the Omicron variant also caused its growth rate to decrease faster, causing changes in the observed growth-rate difference shown earlier (Fig. 2D).

## 4 Discussion

We compare estimates of the forward incubation-period and generation-interval distributions of the Delta and Omicron variants from the Netherlands in late 2021 and early 2022. The original analysis detailing the data set previously reported a shorter mean incubation period and serial interval for the Omicron variant [4]. Accounting for differences in epidemic growth rates, however, we find similar incubation-period distributions for both variants but a shorter (by 0.3–0.8 days) mean generation interval for the Omicron variant relative to that of the Delta variant. Finally, we estimate that the transmission advantage of the Omicron variant decreased from 2.1-fold to 1.4-fold between early December and late January. Improving generation-interval estimates by taking dynamical effects into account may improve understanding of epidemic dynamics and control measures.

The generation-interval distribution describes changes in the individual-level transmission dynamics over the course of infection and therefore provides crucial information for epidemic control. A few studies have estimated the generation-interval distributions of SARS-CoV-2 infections from serial-interval data, but most of them neglect the effects of epidemic growth rates [10, 13, 14, 15]—these practices can be largely attributed to historical work that concluded that serial and generation intervals have the same means based on the assumption that infectors and infectees have identical incubation-period distributions [11, 12, 27]. We build on newer work [3], which demonstrated theoretically that forward serial-interval distributions depend on epidemic growth rates, and further confirm that estimates of the forward generation-interval distributions are indeed sensitive to epidemic growth rates. These effects are also pertinent to epidemiological inferences of past events from a cohort of infected individuals who experienced a later event at the same time—this includes inferences of other delay distributions, such as incubation-period distributions, as well as viral load trajectories [28]. Our sensitivity analysis also shows that the assumptions about the correlation between incubation periods and generation intervals can also have important effects on the estimates of the generation-interval distributions (Supplementary Figure S1).

This study presents a method for accounting for dynamical biases when inferring incubation-period distributions based on epidemic growth rates. Observed incubation-period distributions based on symptom-based cohorts are generally expected to be biased, and similar kinds of corrections will be necessary to accurately estimate the incubation-period distribution. We note that making these kinds of corrections will also depend on data availability, model complexity, and other epidemiological covariates affecting incubation periods, such as vaccine statuses. Accounting for different sources of biases is critical to accurately estimating incubation-period distributions (and other epidemiological distributions alike) but will necessarily increase uncertainties in the estimates. On the other hand, it is still possible to characterize the forward incubation-period distributions without making growth-rate-based corrections through a careful cohorting of individuals with similar infection times when detailed information about infection time is available—we were not able to explore this in our analysis because we relied on publicly available information, which do not contain individual-level information, such as exposure or symptom onset dates.

A few studies have suggested that the incubation period of the Omicron variant may be shorter than that of the Delta variant. The median estimates of the Omicron incubation period typically range between 3–4 days, consistent with earlier findings of [4]. However, these data were collected when the number of Omicron infections was growing rapidly [29, 30], suggesting that they may have been subject to similar biases. On the other hand, incubation-period estimates based on individuals who were exposed from the same event are likely more reliable (because they look forward in time): [31] estimated the median incubation period of the Omicron variant to be 3 days among those who attended the same holiday party (*n* = 117) on 26 November 2021 in Norway. However, we cannot rule out the possibility that some of these attendees were infected prior to the party given that some individuals had COVID-like symptoms prior to the party with at least 96 of the attendees sharing offices; neglecting these factors can lead to underestimation of the mean incubation period. Systematic comparisons of data collection methods and epidemiological contexts are needed to properly assess the differences in incubation period distributions of the Delta and Omicron variants.

A few studies have estimated that the Omicron variant has shorter transmission intervals than the Delta variant [2, 32, 30], but there has been a lack of direct generation-interval estimates. [33, 34] tried to estimate the generation-interval distributions of the Omicron variant but they both relied on population-level epidemic dynamics (rather than individual-level transmission data). Although we estimate a shorter mean generation interval for the Omicron variant, we find the generation-interval distribution of the Omicron and Delta variants have similar modes (around 2.5 days), implying that the realized transmissibility of the Omicron variant decays faster. We tentatively hypothesize that these differences may be primarily driven by the network effect [22, 15]: a higher reproduction number of the Omicron variant leads to faster susceptible depletion among close contacts, which in turn prevents long generation intervals from generating infections. Previous simulations showed that network effects can have considerable impact on realized generation intervals even during the initial exponential growth phase, when susceptible depletion is negligible at the population level [22]. While the network effect is expected to be strongest among household contacts, it is also applicable to other forms of contact structures that involve repeated contacts between the same group of individuals (because only the first infectious contact results in infection). Shorter generation-interval estimates for between-household contacts may be attributable to behavioral effects: individuals who have symptoms or tested positive may be more likely to stay home, preventing long between-household transmission. Other factors, such as more stringent contact tracing measures against the Omicron variant in the Netherlands [4] faster within-host clearance of the Omicron variant [35], and viral kinetics of reinfection and breakthrough infections, also likely contributed to shortening of generation intervals. If shorter generation intervals of the Omicron variant represents an increased proportion of presymptomatic transmission, control measures that target symptomatic individuals can become less effective.

While our study indicates that the Omicron variant has a shorter mean realized generation interval than that of the Delta variant, it is still uncertain how infectiousness profiles differ intrinsically between Omicron and Delta. In particular, similarities in the incubation-period distributions of the Delta and Omicron variants suggest that the differences in their true infectiousness profile may be smaller than the estimated differences in their realized generation-interval distributions. In addition, the “intrinsic” generation intervals of both Omicron and Delta variants are likely longer than what we estimate given existing levels of interventions, including vaccination, and pandemic awareness—estimating intrinsic (or “unmitigated”) generation-interval distributions of SARS-CoV-2 variants is expected to be a difficult problem as it requires data from times when awareness levels were low [19]. Nonetheless, estimates of realized generation-interval distributions describe current epidemic dynamics, implicitly accounting for intervention and behavioral effects, and can therefore be expected to improve estimates of effective reproduction numbers.

Our study also has important implications for estimating transmission advantages of new SARS-CoV-2 variants. In the example we consider, neglecting differences in the generation-interval distributions leads to ≈10% bias in the estimates of the reproduction advantage (i.e., the ratio between the reproduction numbers of the Omicron and Delta variants). More generally, the bias in inferring the reproduction advantage of an emerging variant is expected to be sensitive to the assumed generation-interval distribution of the resident variant. For example, [36] estimated a much higher reproduction advantage of the Omicron variant (> 4-fold) compared to the Delta variant in South Africa but also assumed a longer mean generation interval for the Delta and Omicron variants (6.4 vs 5.2 days, respectively). With our generation-interval estimates, we estimate a 2.6-fold reproduction advantage for the Omicron variant assuming *r* = −0.06 and *r* = 0.26 for the Delta and Omicron variants, respectively—these growth rates were chosen to match the 4-fold reproduction advantage with the previously assumed generation-interval distributions and estimated growth-rate differences of 0.32/day for the Gauteng province, South Africa [36].

We considered two ways of measuring transmission advantages: growth-rate differences and reproduction advantage. Characterizing new variants in terms of their reproduction advantage is useful because it is directly related to the amount of increased transmissibility and immune evasion [36]. On the other hand, the growthrate difference is easier to estimate in real time, and is also more directly relevant to short-term dynamics. For example, when two strains have the same ℛ > 1, the one with shorter generation intervals will grow faster and become dominant as long as ℛ > 1; however, when ℛ is reduced below 1 (either due to intervention or susceptible depletion), the one with longer generation interval will grow faster. These transmission advantages are captured by the growth-rate difference, but not by the ratio of reproduction numbers of two strains. Therefore, we suggest using growth-rate differences and reproduction advantage as complementary measures for understanding the dynamics of emerging SARS-CoV-2 variants.

There are several limitations to our analysis. First, we primarily rely on case data to understand epidemic patterns of the Delta and Omicron variants. In doing so, we implicitly assume that the delay between infection and reports is fixed. However, changes in case trajectories are sensitive to testing patterns and therefore may not accurately reflect patterns of infections. While this limitation does not affect our generation-interval estimates, our inferences of the transmission advantages of the Omicron variant should be interpreted with care.

We assume a constant growth rate for each variant throughout our analysis. During the study period, growth rates of both the Delta and Omicron variants changed slowly, and therefore our constant-growth-rate assumption provides a reasonable approximation for their dynamics across two weeks. However, this assumption might be problematic when growth rates are changing rapidly (e.g., due to an introduction of stringent control measures) or if the sampling window is too wide. Extending our framework to account for time-varying growth rates is relatively simple when inferring the forward incubation-period distribution from the corresponding backward distribution—we can simply replace *r* with *r*(*t*) in Eq. (2) because the backward incubation-period distribution is a weighted average of the forward incubation-period distributions and the number of individuals in each cohort (i.e., individuals who were infected at the same time). However, such extensions are more complicated for linking generation- and serial-interval distributions because the forward serial-interval distribution also depends on the cohort reproduction number—for example, if a certain cohort of infectors had higher reproduction number (e.g., because they were infected before control measures were observed), we are more likely to observe transmission from this cohort (see [3] for more details). Assuming exponential growth allows us to avoid this complexity. Extending our framework to account for timevarying growth rates can provide more accurate tools for inferring epidemiological delay distributions.

We do not account for individual-level heterogeneity, such as age, vaccination status, or previous exposure history. In general, epidemic growth rates may differ between infection groups (e.g., the incidence of infection caused by any variant is expected to grow faster among immunologically naive individuals), and these growth-rate differences can affect estimates of epidemiological delay distributions, including the incubation-period and generation-interval distributions. We are not able to perform stratified analyses because individual-level information was not publicly available. Therefore, while we estimate unclear differences in incubation-period distributions between Delta and Omicron infections, controlling for other covariates, such as age and immune history, may help better characterize differences in Delta and Omicron infections.

Finally, there are several sources of biases in serial-interval data that we did not consider. For example, the direction of transmission is difficult to establish for SARS-CoV-2 due to pre-symptomatic transmission. Other sources of information, such as exposure history and positive test results, can help resolve uncertainties but are imperfect. Serial-interval data also depend on the ability of infected individuals to accurately recall when their symptoms started. Future studies may explore how these biases affect the inference of generation intervals from serial intervals. While comparisons of incubation-period and serial-interval distributions can shed insight on pathogen dynamics, both distributions typically do not account for the dynamics of asymptomatic infections; neglecting these differences can further bias estimates of transmissibility of a pathogen [37].

Monitoring changes in key epidemiological parameters is critical to understanding the evolution of SARS-CoV-2 and predicting its future dynamics [38]. Our study synthesizes a previously developed theoretical framework on serial- and generation-interval distributions and presents methodological advances in monitoring epidemiological parameters. Similar efforts will be critical to improve estimates of the infectiousness profiles of future SARS-CoV-2 variants, especially among asymptomatically infected individuals. These conclusions are relevant for other emerging and endemic pathogens in general.

## Data availability

All data and code are stored in a publicly available GitHub repository (https://github.com/parksw3/omicron-generation).

## Acknowledgements

We thank Ron Milo for providing helpful comments on the manuscript. JD was supported by the Canadian Institutes of Health Research, the Natural Sciences and Engineering Research Council of Canada, and the Michael G. DeGroote Institute for Infectious Disease Research. JSW acknowledges support from the Army Research Office W911NF1910384 and the Ile de France region via the Chaires Blaise Pascal program. JW and JB has received funding from European Union’s Horizon 2020 research and innovation programme—project EpiPose, Epidemic intelligence to Minimize COVID-19’s Public Health, Societal and Economical Impact (grant agreement number 101003688). SF was supported by the Wellcome Trust (grant no. 210758/Z/18/Z). The funders had no role in study design, data collection and analysis, decision to publish, or preparation of the manuscript. The findings and conclusions in this study are those of the authors and do not necessarily represent the official position of the funding agencies, the National Institutes of Health, or the U.S. Department of Health and Human Services.

## Supplementary Materials

**Supplementary Figure S1:**
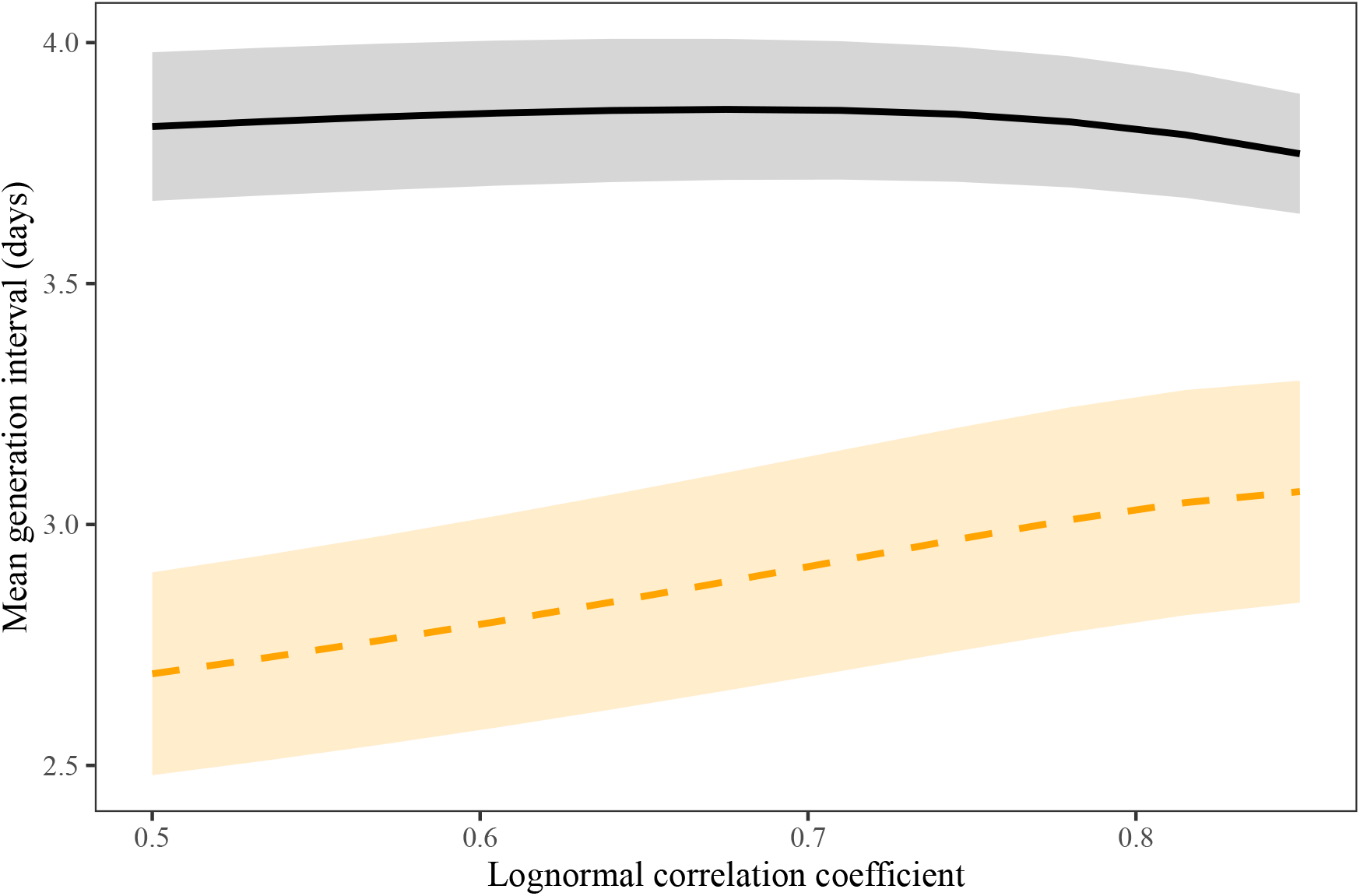
Sensitivity of the estimates of the mean generation interval to the assumed values of the correlation coefficient of the lognormal distribution. Lines and shaded regions represent maximum likelihood estimates and the corresponding 95% confidence intervals for the Delta (black, solid lines) and Omicron variants (orange, dashed lines). For illustrative purposes we use within-household serial-interval data from the cohort of infectors who developed symptoms during weeks 50 (13–19 December) and 51 (20–26 December) of 2021.

**Supplementary Figure S2:**
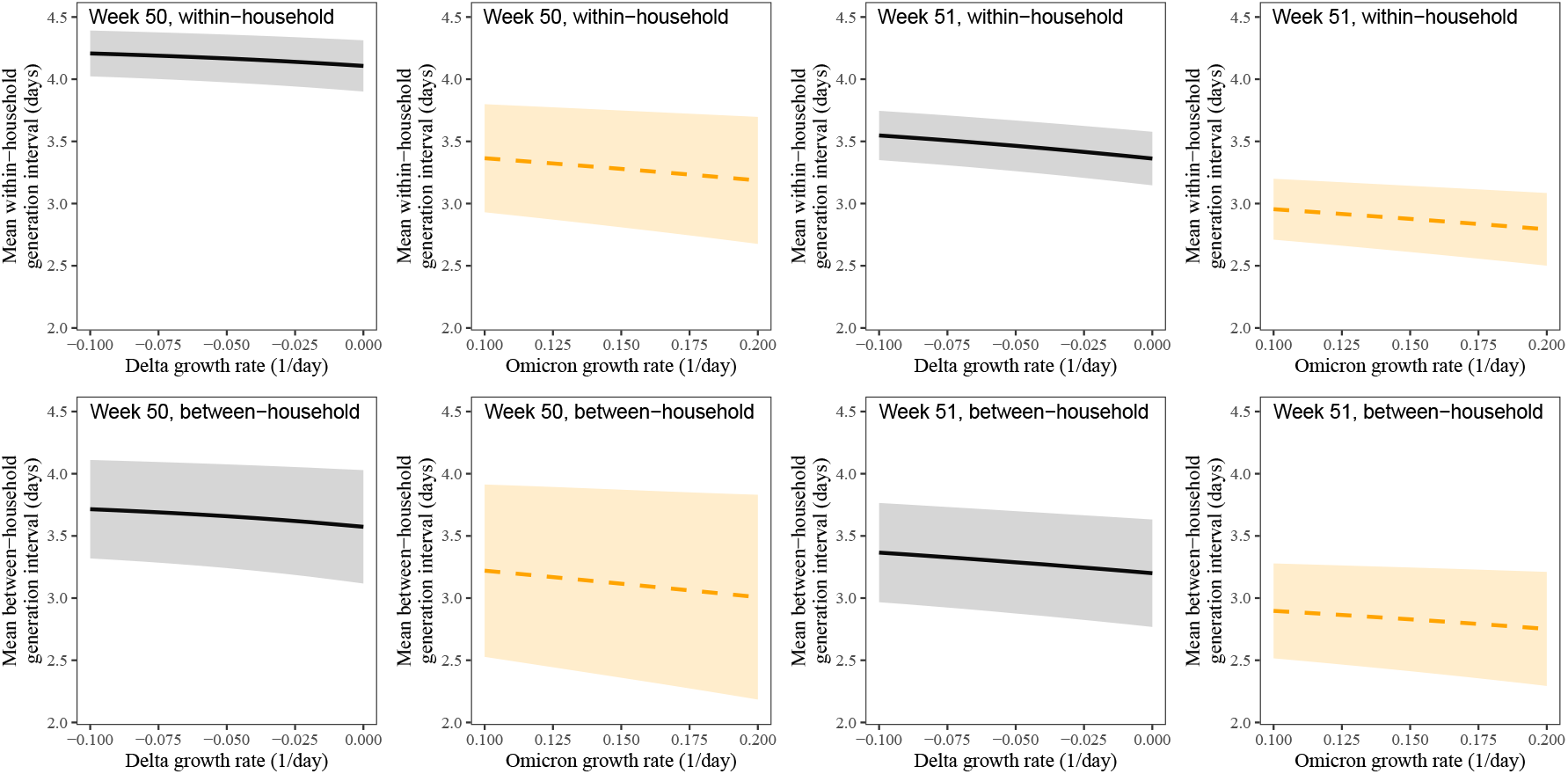
Estimated mean forward generation intervals of Delta and Omicron variants across different stratifications. Sensitivity of the mean forward generation-interval estimates to assumed growth rates of the Delta and Omicron variants stratified by the types of transmission (within- vs between-household transmission) and the week of infectors’ symptom onset (week 50, 13–19 December 2021, vs week 51, 20–26 December 2021,). Lines represent maximum likelihood estimates. Shaded regions represent the corresponding 95% confidence intervals.

**Supplementary Figure S3:**
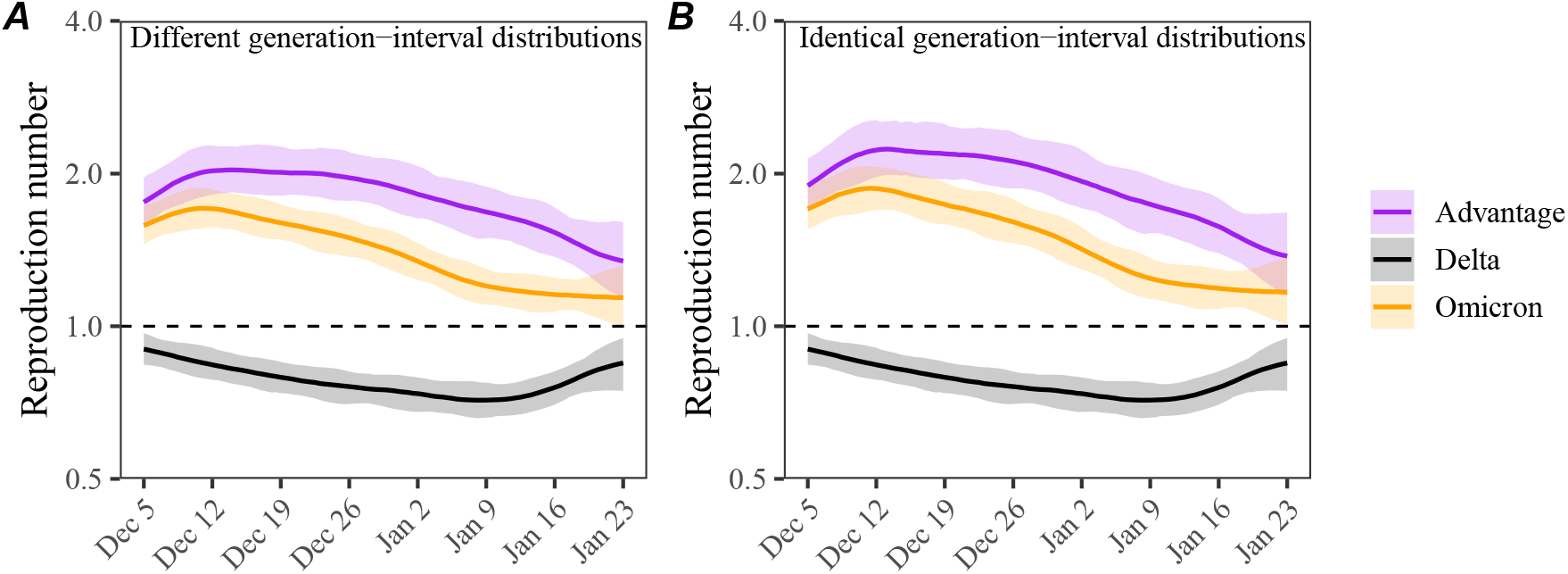
Estimated time-varying reproduction number advantages of the Omicron variant using between-household generation-interval distributions. (A) Estimated instantaneous reproduction numbers and their ratios over time while accounting for differences in the generation-interval distributions. (B) Estimated instantaneous reproduction numbers and their ratios over time while assuming identical generation-interval distributions. The instantaneous reproduction number of each variant is estimated using the renewal equation by shifting the smoothed case curves by one week (Fig. 2C). The intrinsic generation-interval distribution is approximated by the maximum likelihood estimates of the forward generation-interval distributions for between-household transmission pairs based on *r* = −0.05 for the Delta variant (black) and *r* = 0.15 for the Omicron variant (orange). Purple lines represent the ratio between the effective reproduction numbers of the Delta and Omicron variants. Lines and shaded regions represent medians and corresponding 95% confidence intervals.

